# Factors Influencing Knowledge and Utilization of Insecticide-Treated Nets among Undergraduate Hostel Residents at the University of Port Harcourt, Nigeria

**DOI:** 10.1101/2025.11.23.25340790

**Authors:** Harrison Oghenevwegba Odibo, Lauretta Oluchi Ukor, Similoluwa Dolapo Adeleye, Fortune Ita Sunday, Wilcox Adonye Shekinah, Emmanuel Chimzi Weje

**Affiliations:** University of Calabar, Calabar, Cross River State, Nigeria; University of Port Harcourt, Port Harcourt, Rivers State, Nigeria; Edo State University, Uzairue, Edo State, Nigeria; University of Uyo, Uyo, Akwa Ibom State, Nigeria

**Author notes:** Corresponding Author: Harrison Oghenevwegba Odibo, University of Calabar, PMB 1115, Calabar, Cross River State, Nigeria. Email: [ ].

**Keywords:** Malaria, Insecticide-Treated Nets, Knowledge, Utilization, University Students, Nigeria, Prevention

## Abstract

**Background:** Malaria remains a significant public health burden in Nigeria. Insecticide-treated nets (ITNs) are a proven effective intervention, yet their utilization, particularly among understudied populations like university students, is often suboptimal. This study assessed the knowledge, prevalence of use, and factors associated with ITN utilization among undergraduate hostel residents at the University of Port Harcourt.

**Methods:** A descriptive cross-sectional study was conducted among 374 undergraduate hostel residents selected via a multi-stage sampling technique. Data were collected using a pre-tested, self-administered questionnaire. Analysis was performed using IBM SPSS version 26, employing descriptive statistics and chi-square tests at a 5% significance level.

**Results:** The mean age of respondents was 20.1 ± 2.3 years, with a female predominance (58.6%). Knowledge of ITNs was high, with 100% having heard of ITNs and 46.5% possessing good knowledge. Health workers (61.5%) were the primary source of information. However, only 33.4% reported using ITNs every night. Significant factors associated with ITN use included female gender (p=0.016), residence in the Abuja campus (p=0.002), and enrollment in the Health Sciences faculty (p=0.005). The most common barriers to use were discomfort from heat (48.8%) and itchiness (48.8%) among users who reported adverse effects.

**Conclusion:** A significant gap exists between high knowledge and low practice of ITN use among students. The primary deterrents were user discomfort and adverse effects. To bridge this gap, interventions should focus on distributing more comfortable, user-friendly nets and implementing targeted behavioral change communication, especially for male students and non-health science faculties, to translate knowledge into consistent practice.

## 1. Introduction

Malaria persists as a major cause of morbidity and mortality in tropical regions. In 2022, there were an estimated 249 million cases globally, with the WHO African Region bearing the heaviest burden, accounting for 94% of cases and 95% of deaths [1]. Nigeria is the most affected country, contributing to nearly 27% of the global malaria burden [1].

The World Health Organization (WHO) recommends insecticide-treated nets (ITNs) as a cornerstone of malaria prevention. ITNs are estimated to be twice as effective as untreated nets and have been shown to reduce child mortality and uncomplicated malaria episodes significantly [2, 3]. Consequently, they are central to malaria control programs worldwide, including Nigeria’s National Malaria Elimination Programme (NMEP) [4].

Extensive research has documented the knowledge and utilization of ITNs among traditional vulnerable groups such as pregnant women and children under five. Studies among pregnant women in South-Eastern Nigeria and antenatal clinic attendees in Abuja found high levels of knowledge (over 90%) about ITNs [5, 6]. However, this high knowledge does not consistently translate to practice. For instance, while a study in Lagos State found strong awareness, actual usage was as low as 11.2% in some areas, primarily due to discomfort and heat [7]. Similar knowledge-practice gaps have been observed in rural Southwestern Uganda and Northwest Ethiopia, where barriers like perceived discomfort, misconceptions about chemicals, and difficulty in hanging nets were prevalent [8, 9].

The university student population, particularly those in hostel accommodations, represents a unique and understudied demographic. Hostel environments, characterized by high occupant density and potential for poor sanitation, can be conducive to mosquito breeding and malaria transmission [10]. Despite this risk, students are often neglected in mass ITN distribution campaigns, which typically target households with pregnant women and young children. A previous study at the University of Port Harcourt by Douglas et al. confirmed a high awareness of ITNs among undergraduates but noted a poor corresponding attitude and practice [11]. This suggests that the factors influencing ITN use in this specific demographic may be distinct and require further investigation.

Theoretical models like the Health Belief Model (HBM) provide a useful framework for understanding this gap between knowledge and action [12]. The HBM posits that health behavior is influenced by an individual’s perception of susceptibility to a disease, the severity of the disease, the benefits of an action, and the barriers to taking that action. In the context of ITN use, students may perceive a low susceptibility to severe malaria or perceive barriers (discomfort, heat) that outweigh the benefits.

Therefore, building on the preliminary findings of Douglas et al. [11], this study aimed to conduct a deeper investigation into the factors affecting the knowledge and, crucially, the use of ITNs among undergraduate hostel residents at the University of Port Harcourt. By identifying the specific barriers and facilitators within this community, the findings will inform the development of targeted interventions to improve ITN utilization and reduce the burden of malaria in this setting.

## 2. Methods

### 2.1 Study Design and Area

A descriptive cross-sectional study was conducted in June 2023 at the University of Port Harcourt, a federal university in Rivers State, Nigeria. The university has over 40,000 students, with approximately 3,000 residing in 24 hostels across its Abuja, Delta, and Choba campuses.

### 2.2 Study Population and Sampling

The study population was undergraduate students residing in university hostels. The sample size was calculated as 374 using the formula for a single proportion, with a 33% prevalence of ITN use from a previous study [11], a 95% confidence level, and a 5% margin of error, with a 10% non-response adjustment.

A multi-stage sampling technique was employed:

- **Stage 1:** Two campuses (Abuja and Delta) were selected by simple random sampling (balloting).
- **Stage 2:** Ten hostels (five from each selected campus) were selected by simple random sampling.
- **Stage 3:** One floor was randomly selected from each hostel, and 38 respondents were recruited consecutively from rooms on that floor until the sample size was attained.

### 2.3 Data Collection and Instrument

Data were collected using a structured, self-administered questionnaire, divided into four sections:

- Section A: Socio-demographic characteristics.
- Section B: Knowledge of ITNs (15 items, scored; poor: <50%, moderate: 50-75%, good: >75%).
- Section C: Attitudes towards ITNs.
- Section D: ITN use practices and barriers.

The questionnaire was pre-tested and validated for clarity and consistency.

### 2.4 Data Analysis

Data were analyzed using IBM SPSS Statistics version 26. Categorical variables were summarized using frequencies and percentages. The association between categorical variables (e.g., socio-demographics and ITN use) was assessed using the Chi-square test, with a p-value of <0.05 considered statistically significant.

### 2.5 Ethical Consideration

Ethical approval was obtained from the Department of Preventive and Social Medicine, University of Port Harcourt. Written informed consent was obtained from all participants before questionnaire administration. Anonymity and confidentiality were maintained throughout the study.

## 3. Results

### 3.1 Socio-demographic Characteristics

A total of 374 students participated, yielding a 100% response rate. The mean age was 20.1 (±2.3) years, with the largest proportion (45.7%) aged 15-19. There were more females (58.6%) than males. The majority were single (98.7%), Christian (96.3%), and first-year students (33.2%). Most respondents (79.9%) resided in the Abuja campus, and 24.9% were from the College of Health Sciences (Table 1).

**Table 1.**
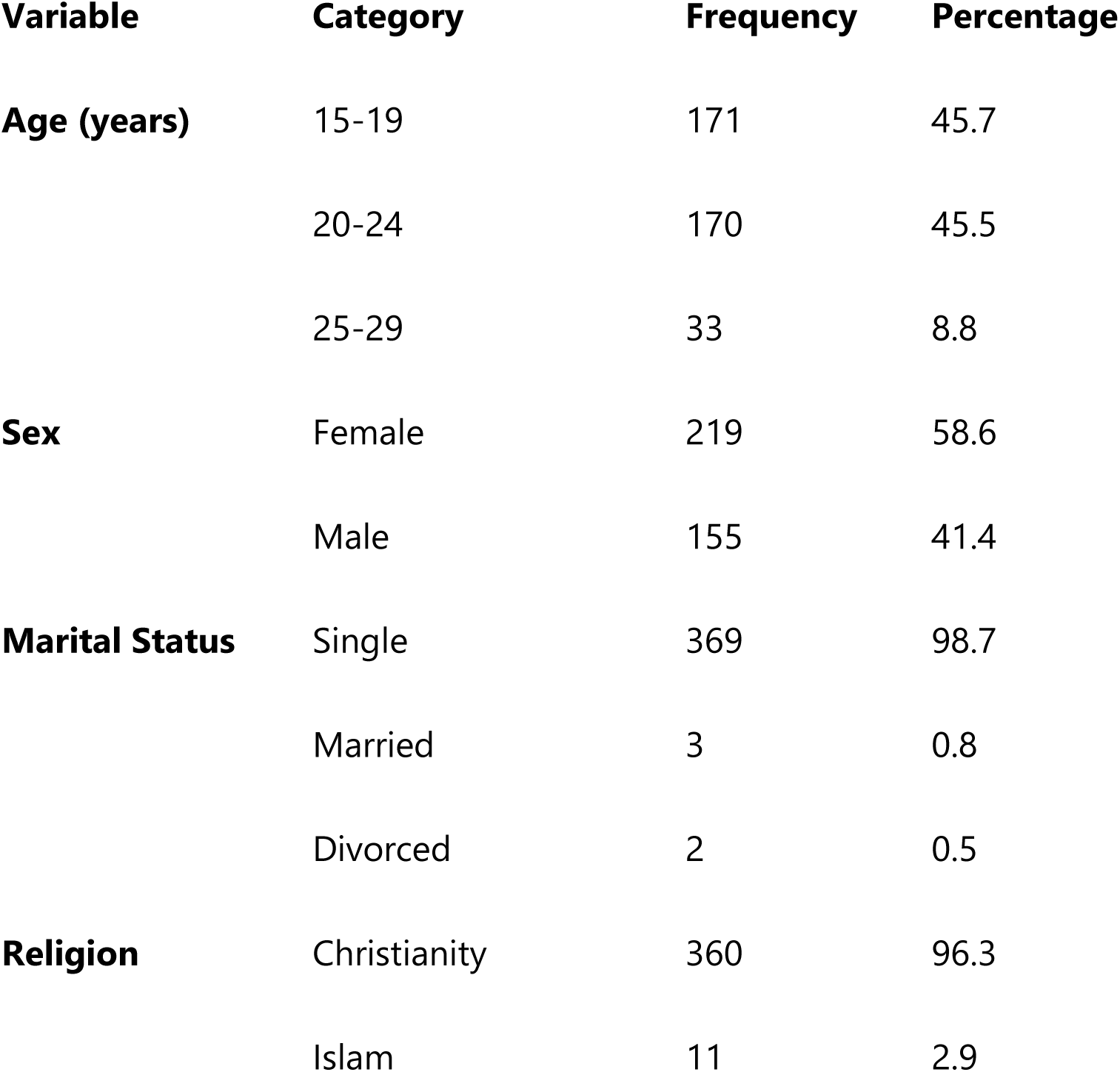
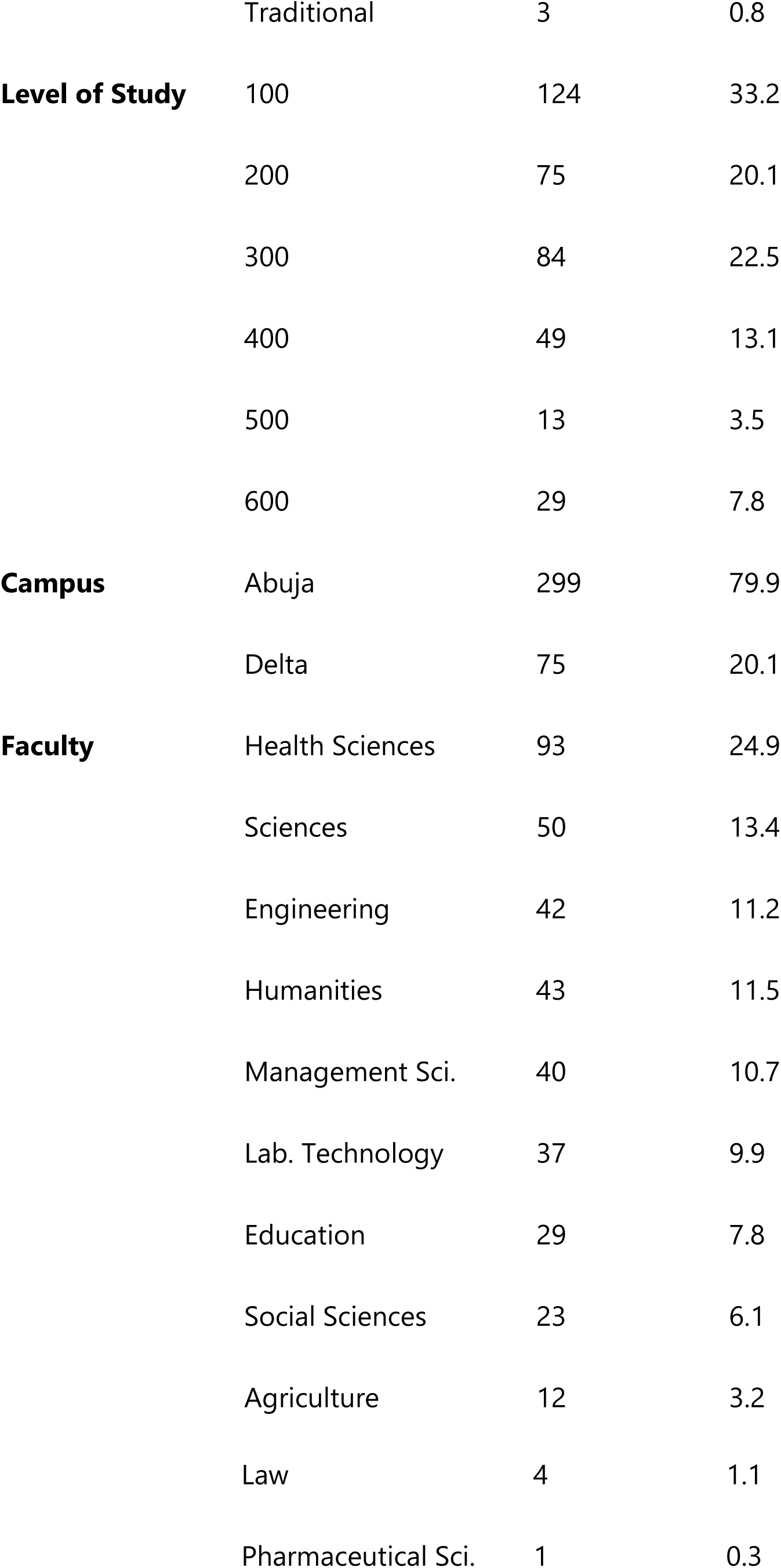
Socio-demographic Characteristics of Respondents (n=374)

### 3.2 Knowledge of Insecticide-Treated Nets

All respondents (100%) had heard of ITNs, and 87.7% had seen or handled one. Nearly all (98.4%) correctly identified ITNs as a tool for preventing mosquito bites/malaria. Health workers (61.5%) were the most common source of information. The level of knowledge was categorized as good in 46.5% of respondents, moderate in 52.4%, and poor in only 1.1%. Good knowledge was significantly associated with the faculty of study (p=0.003), being highest among Health Sciences students.

### 3.3 Attitude and Prevalence of ITN Use

Although 57.8% of respondents owned an ITN and 93.9% had used one at least once, only 33.4% (125/374) reported using it every night, while 66.6% (249/374) did not. A majority (86.1%) stated they would use an ITN if it were free. The most common trigger for use was having doors and windows open (66.8%). ITN use was significantly higher among females (67.2%, p=0.016), students in the Abuja campus (88.8%, p=0.002), and those in the Health Sciences faculty (36.0%, p=0.005) (Table 2).

**Table 2.**
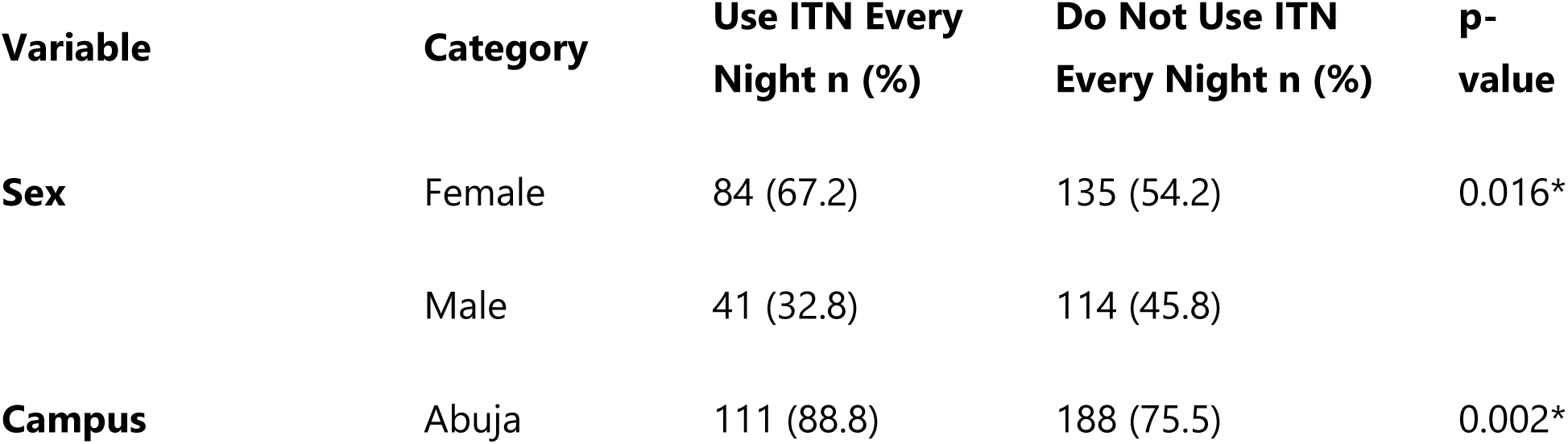
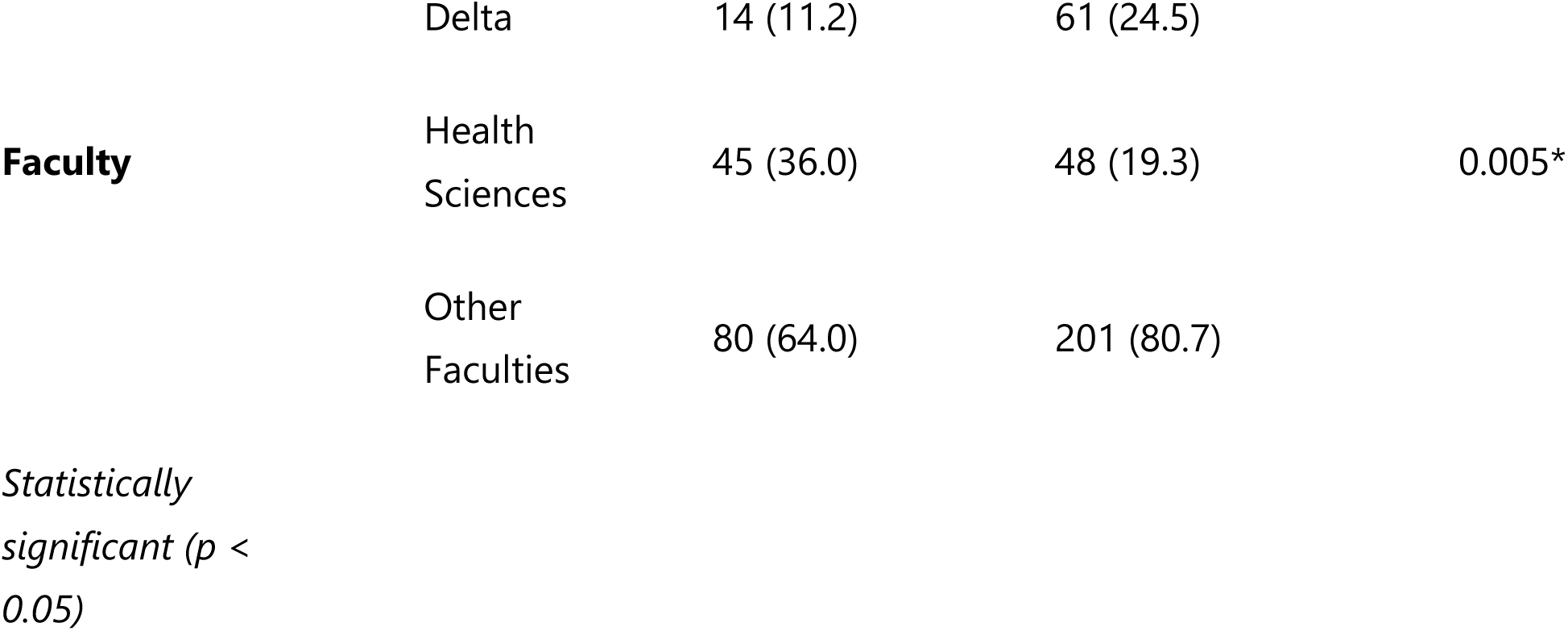
Factors Associated with Consistent ITN Use (n=374)

### 3.4 Practices and Adverse Effects Associated with ITN Use

Among users, 30.5% obtained their nets from health workers. Of the 351 respondents who had ever used an ITN, 248 (70.7%) reported experiencing at least one adverse effect. The most reported adverse effects were itchiness (48.8%) and increased heat (48.8%). Other effects included rashes (21.8%), catarrh (12.9%), and headache (6.5%) (Table 3).

**Table 3.** Adverse Effects Reported by ITN Users (n=248)

| Adverse Effect | Frequency | Percentage |
| --- | --- | --- |
| Increased Heat | 121 | 48.8 |
| Itchiness | 121 | 48.8 |
| Rashes | 54 | 21.8 |
| Catarrh | 32 | 12.9 |
| Headache | 16 | 6.5 |
| Dizziness | 10 | 4.0 |
| Others | 21 | 8.5 |

*Note: Multiple responses were possible.*

## 4. Discussion

This study reveals a critical gap between knowledge and practice regarding ITN use among undergraduate students at the University of Port Harcourt, a finding consistent with research among other demographics in Nigeria and sub-Saharan Africa [5, 7, 8]. While knowledge was high, consistent usage was low, underscoring that knowledge is a necessary but insufficient condition for behavioral change.

The high level of ITN knowledge aligns with studies conducted among university staff in Rivers State [13] and students in Ghana [14], and can be attributed to sustained public health campaigns. The finding that health workers were the primary source of information reinforces their critical role as health educators beyond clinical settings [13]. The significantly better knowledge among Health Sciences students was expected and mirrors findings by Douglas et al. [11], highlighting the impact of specialized education on health literacy.

The prevalence of consistent ITN use (33.4%) is discouragingly low but comparable to the 31.6% found among male students at the University of Lagos [15] and the low utilization despite high awareness reported by Anikwe et al. in Ebonyi State [16]. This pervasive knowledge-practice gap can be effectively analyzed through the lens of the Health Belief Model [12]. While students likely perceive the severity of malaria, our findings suggest that **perceived barriers**—primarily physical discomfort (heat and itchiness)—strongly outweigh the perceived benefits of using an ITN. This resonates with studies that identify discomfort as a principal barrier across diverse populations [7, 8, 9].

The significant association between ITN use and female gender corroborates global trends showing women often exhibit more proactive health-seeking behaviors [17]. The higher usage among Health Sciences students further emphasizes that deeper knowledge, likely coupled with a higher perceived susceptibility from clinical exposure, can positively influence practice. The reason for higher usage in the Abuja campus is unclear and warrants further qualitative investigation into potential peer influence or infrastructural differences.

The high prevalence of adverse effects, particularly heat and itchiness, is a major public health concern and a direct explanation for the low usage. These findings are consistent with reports of skin irritation and discomfort from other studies [8, 18]. This underscores an urgent need for the development and distribution of “next-generation” ITNs made with more comfortable fabrics and insecticides with lower dermal and respiratory irritation profiles.

### 4.1 Limitations of the Study

This study relied on self-reported data, which is subject to social desirability bias. The cross-sectional design establishes associations but not causality. Furthermore, the study was conducted in one university, which may limit the generalizability of the findings to all Nigerian universities.

## 5. Conclusion and Recommendations

This study concludes that a significant knowledge-practice gap regarding ITN use exists among students at the University of Port Harcourt. The primary impediments are perceived barriers related to user discomfort, chiefly heat and itchiness.

To address this, the following recommendations are proposed:

- **To University Authorities:**

1. Partner with public health agencies to distribute **“comfort-optimized” ITNs** and integrate **practical demonstrations** on ITN use into orientation programs.
2. Improve hostel infrastructure, such as ensuring **adequate power supply** for fans, to mitigate the discomfort of heat.
- **To Public Health Agencies & Government:**

1. Invest in **research and development** of next-generation ITNs that minimize heat retention and skin irritation.
2. Design **targeted behavioral change communication** for youths, addressing the specific barriers of discomfort.
- **To Students:**

1. Student bodies should organize **peer-led awareness campaigns** and create channels for **feedback** on distributed ITNs.

## Data Availability

All data produced in the present study are available upon reasonable request to the authors

---

**Table S1.** SOCIO-DEMOGRAPHIC CHARACTERISTICS OF STUDY PARTICIPANTS.

| **Variables** | **Frequency** | **Percentage** |
| --- | --- | --- |
| **Age** |  |  |
| 15-19 years | 171 | 45.7 |
| 20-24 years | 170 | 45.5 |
| 25-29 years | 33 | 8.8 |
| **Sex** |  |  |
| Female | 219 | 58.6 |
| Male | 155 | 41.4 |
| **Marital Status** |  |  |
| Divorced | 2 | 0.5 |
| Married | 3 | 0.8 |
| Single | 369 | 98.7 |
| **Religion** |  |  |
| Christianity | 360 | 96.3 |
| Islam | 11 | 2.9 |
| Traditional | 3 | 0.8 |
| **Level** |  |  |
| 100 | 124 | 33.2 |
| 200 | 75 | 20.1 |
| 300 | 84 | 22.5 |
| 400 | 49 | 13.1 |
| 500 | 13 | 3.5 |
| 600 | 29 | 7.8 |
| **Campus** |  |  |
| Abuja | 299 | 79.9 |
| Delta | 75 | 20.1 |
| **Faculty of study** |  |  |
| Agriculture | 12 | 3.2 |
| Education | 29 | 7.8 |
| Engineering | 42 | 11.2 |
| Faculty of Law | 4 | 1.1 |
| Health sciences | 93 | 24.9 |
| Humanities | 43 | 11.5 |
| Laboratory Technology | 37 | 9.9 |
| Management sciences | 40 | 10.7 |
| Pharmaceutical science | 1 | 0.3 |
| Sciences | 50 | 13.4 |
| Social sciences | 23 | 6.1 |

**Table S2.** KNOWLEDGE OF INSECTICIDE TREATED NETS.

| **Variables** | **Frequency** | **Percentage** |
| --- | --- | --- |
| **Heard of ITN** |  |  |
| No | 0 | 0.0 |
| Yes | 374 | 100.0 |
| **Seen or Handled ITN** |  |  |
| No | 46 | 12.3 |
| Yes | 328 | 87.7 |
| **Uses of ITN** |  |  |
| Beautification | 2 | 0.5 |
| Prevent Mosquito | 368 | 98.4 |
| Sound sleep | 4 | 1.1 |
| **Where can ITN be obtained** |  |  |
| **Health center** |  |  |
| No | 22 | 5.9 |
| Yes | 352 | 94.1 |
| **Chemical stores** |  |  |
| No | 317 | 84.8 |
| Yes | 57 | 15.2 |
| **Market** |  |  |
| No | 267 | 71.4 |
| Yes | 107 | 28.6 |
| **Others** |  |  |
| No | 366 | 97.9 |
| Yes | 8 | 2.1 |
| **What should be done before the first usage of ITN** |  |  |
| Air it | 319 | 85.3 |
| Chemical addition | 8 | 2.1 |
| Wash it | 43 | 11.5 |
| Others | 4 | 1.1 |
| **Can ITN be retreated** |  |  |
| No | 39 | 10.4 |
| Yes | 335 | 89.6 |
| **When should ITN be treated** |  |  |
| After 3-5 years of usage | 158 | 42.2 |
| After washing three times | 59 | 15.8 |
| After washing once | 29 | 7.8 |
| After you had a bite while using | 74 | 19.8 |
| When it is dirty | 54 | 14.4 |
| **How long can ITN effectiveness last** |  |  |
| 1-7 days | 17 | 4.5 |
| Weeks | 32 | 8.6 |
| 1-3 months | 90 | 24.1 |
| 4-12 months | 61 | 16.3 |
| 1-2 years | 67 | 17.9 |
| 3-5 years | 107 | 28.6 |
| **Sources of information for ITNs** |  |  |
| **Health workers** |  |  |
| No | 144 | 38.5 |
| Yes | 230 | 61.5 |
| **Clinic** |  |  |
| No | 222 | 59.4 |
| Yes | 152 | 40.6 |
| **Family and friends** |  |  |
| No | 206 | 55.1 |
| Yes | 168 | 44.9 |
| **Television/Radio/Mass Media** |  |  |
| No | 225 | 60.2 |
| Yes | 149 | 39.8 |
| **Others** |  |  |
| No | 359 | 96.0 |
| Yes | 15 | 4.0 |

**Table S3.** ATTITUDE TOWARDS USE OF INSECTICIDE TREATED NETS BY STUDY PARTICIPANTS.

| **Variables** | **Frequency** | **Percentage** |
| --- | --- | --- |
| **Do you have an ITN** |  |  |
| No | 158 | 42.4 |
| Yes | 216 | 57.8 |
| **Been a beneficiary of ITN** |  |  |
| No | 198 | 52.9 |
| Yes | 176 | 47.1 |
| **Have you ever used ITN** |  |  |
| No | 23 | 6.1 |
| Yes | 351 | 93.9 |
| **Would you use ITN if it was free** |  |  |
| No | 52 | 13.9 |
| Yes | 322 | 86.1 |
| **Would you pay for ITN** |  |  |
| No | 171 | 45.7 |
| Yes | 203 | 54.3 |
| **Do you sleep under ITN often** |  |  |
| No | 223 | 59.6 |
| Yes | 151 | 40.4 |
| **Did you sleep under ITN last night** |  |  |
| No | 278 | 74.3 |
| Yes | 96 | 25.7 |
| **Which season do you mostly use ITN** |  |  |
| All year round | 150 | 40.1 |
| During dry season | 20 | 5.3 |
| During mosquito season | 87 | 23.3 |
| During raining season | 117 | 31.3 |
| **When are you most likely to use ITN** |  |  |
| After it rains | 61 | 16.3 |
| Doors and windows are opened | 250 | 66.8 |
| In the village | 14 | 3.7 |
| With electricity | 37 | 9.9 |
| Others | 12 | 3.2 |

**Table S4.** USE OF ITN BY STUDY PARTICIPANTS.

| **Variables** | **Frequency** | **Percentage** |
| --- | --- | --- |
| **Do you use ITN every night** |  |  |
| No | 249 | 66.5 |
| Yes | 125 | 33.4 |
| **Had negative experience while using it** |  |  |
| No | 126 | 33.7 |
| Yes | 248 | 66.3 |
| **Do you clean you ITN** |  |  |
| No | 63 | 16.8 |
| Yes | 311 | 83.2 |
| **How often do you clean your ITN** |  |  |
| After one year | 46 | 12.3 |
| I don't know | 63 | 16.8 |
| One month | 126 | 33.7 |
| Six months | 53 | 14.2 |
| Three months | 86 | 23.0 |
| **How did you get your ITN** |  |  |
| Gift | 50 | 13.4 |
| Health worker | 114 | 30.5 |
| Hospital | 80 | 21.4 |
| Market | 69 | 18.4 |
| Ministry | 27 | 7.2 |
| Others | 32 | 8.6 |
| Nil | 2 | 0.5 |
| **How do you use ITN** |  |  |
| Cover self | 6 | 1.6 |
| Fixed to the windows | 14 | 3.7 |
| Hang over bed | 340 | 90.9 |
| Pinned to the door | 12 | 3.2 |
| Nil | 2 | 0.5 |

**Table S5.** ADVERSE EFFECTS ASSOCIATED WITH THE USE OF ITNS.

| **Variables** | **Frequency** | **Percentage** |
| --- | --- | --- |
| **Increased heat (n=248)** |  |  |
| No | 127 | 51.2 |
| Yes | 121 | 48.8 |
| **Itchiness (n=248)** |  |  |
| No | 127 | 51.2 |
| Yes | 121 | 48.8 |
| **Headache (n=248)** |  |  |
| No | 232 | 93.5 |
| Yes | 16 | 6.5 |
| **Dizziness (n=248)** |  |  |
| No | 238 | 96.0 |
| Yes | 10 | 4.0 |
| **Catarrh (n=248)** |  |  |
| No | 216 | 87.1 |
| Yes | 32 | 12.9 |
| **Rashes (n=248)** |  |  |
| No | 194 | 78.2 |
| Yes | 54 | 21.8 |
| **Others (n=248)** |  |  |
| No | 227 | 91.5 |
| Yes | 21 | 8.5 |

**Fig 1.**
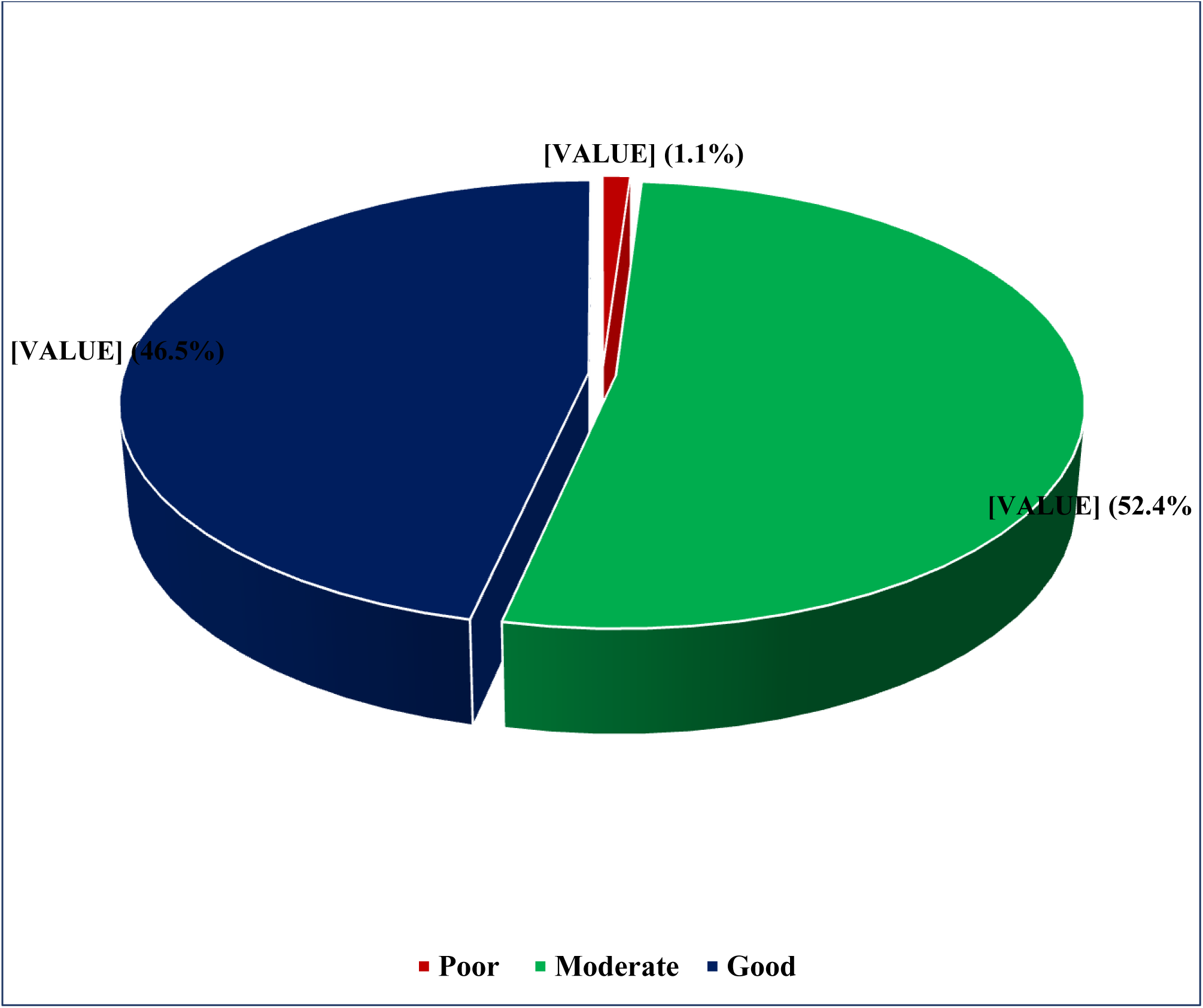
Level of knowledge of ITN among study participants.

**Table S6.** RELATIONSHIP BETWEEN SOCIODEMOGRAPHIC FACTORS AND LEVEL OF KNOWLEDGE OF ITN AMONG UNDERGRADUATES.

| Variables | Level of Knowledge<br>n (%) |  |  | Df | Test<br>Statistics | p-value |
| --- | --- | --- | --- | --- | --- | --- |
|  | Poor<br>n (%) | Average<br>n (%) | Good<br>n (%) |  |  |  |
| **Age** |  |  |  |  |  |  |
| 15-19 years | 3 (75.0) | 95 (48.5) | 73 (42.0) | 4 | $\chi^2=5.974$ | 0.187 |
| 20-24 years | 0 (0.0) | 83 (42.3) | 87 (50.0) |  |  |  |
| 25-29 years | 1 (25.0) | 18 (9.2) | 14 (8.0) |  |  |  |
| **Sex** |  |  |  |  |  |  |
| Female | 1 (25.0) | 117 (59.7) | 101 (58.0) | 2 | $\chi^2=1.979$ | 0.372 |
| Male | 3 (75.0) | 79 (40.3) | 73 (42.0) |  |  |  |
| **Marital Status** |  |  |  |  |  |  |
| Divorced | 0 (0.0) | 2 (10.0) | 0 (0.0) | 4 | $\chi^2=4.602$ | 0.331 |
| Married | 0 (0.0) | 3 (1.5) | 0 (0.0) |  |  |  |
| Single | 4 (100.0) | 191 (97.4) | 174 (100.0) |  |  |  |
| **Religion** |  |  |  |  |  |  |
| Christianity | 4 (100.0) | 189 (96.4) | 167 (96.0) | 4 | $\chi^2=0.640$ | 0.959 |
| Islam | 0 (0.0) | 5 (2.6) | 6 (3.4) |  |  |  |
| Traditional | 0 (0.0) | 2 (1.0) | 1 (0.6) |  |  |  |
| **Level** |  |  |  |  |  |  |
| 100 | 2 (50.0) | 76 (38.3) | 46 (26.4) | 10 | $\chi^2=14.400$ | 0.156 |
| 200 | 0 (0.0) | 37 (18.9) | 38 (21.8) |  |  |  |
| 300 | 1 (25.0) | 41 (20.9) | 42 (24.1) |  |  |  |
| 400 | 0 (0.0) | 24 (12.2) | 25 (14.4) |  |  |  |
| 500 | 1 (25.0) | 6 (3.1) | 6 (3.4) |  |  |  |
| 600 | 0 (0.0) | 12 (6.1) | 17 (9.8) |  |  |  |
| **Campus** |  |  |  |  |  |  |
| Abuja | 4 (100.0) | 155 (79.1) | 140 (80.5) | 4 | $\chi^2=1.123$ | 0.570 |
| Delta | 0 (0.0) | 41 (20.9) | 34 (19.5) |  |  |  |
| **Faculty of study** |  |  |  |  |  |  |
| Agriculture | 0 (0.0) | 8 (4.1) | 4 (2.3) | 20 | $\chi^2=41.499$ | 0.003* |
| Education | 0 (0.0) | 16 (8.2) | 13 (7.5) |  |  |  |
| Engineering | 0 (0.0) | 15 (7.7) | 27 (15.5) |  |  |  |
| Faculty of Law | 1 (25.0) | 2 (1.0) | 1 (0.6) |  |  |  |
| Health sciences | 1 (25.0) | 40 (20.4) | 52 (29.9) |  |  |  |
| Humanities | 1 (25.0) | 28 (14.3) | 14 (8.0) |  |  |  |
| Laboratory Technology | 1 (25.0) | 20 (10.2) | 16 (9.2) |  |  |  |
| Management sciences | 0 (0.0) | 23 (11.7) | 17 (9.8) |  |  |  |
| Pharmaceutical science | 0 (0.0) | 0 (0.0) | 1 (9.8) |  |  |  |
| Sciences | 0 (0.0) | 29 (14.8) | 21 (12.1) |  |  |  |
| Social sciences | 0 (0.0) | 15 (7.7) | 8 (4.6) |  |  |  |

**Table S7.**
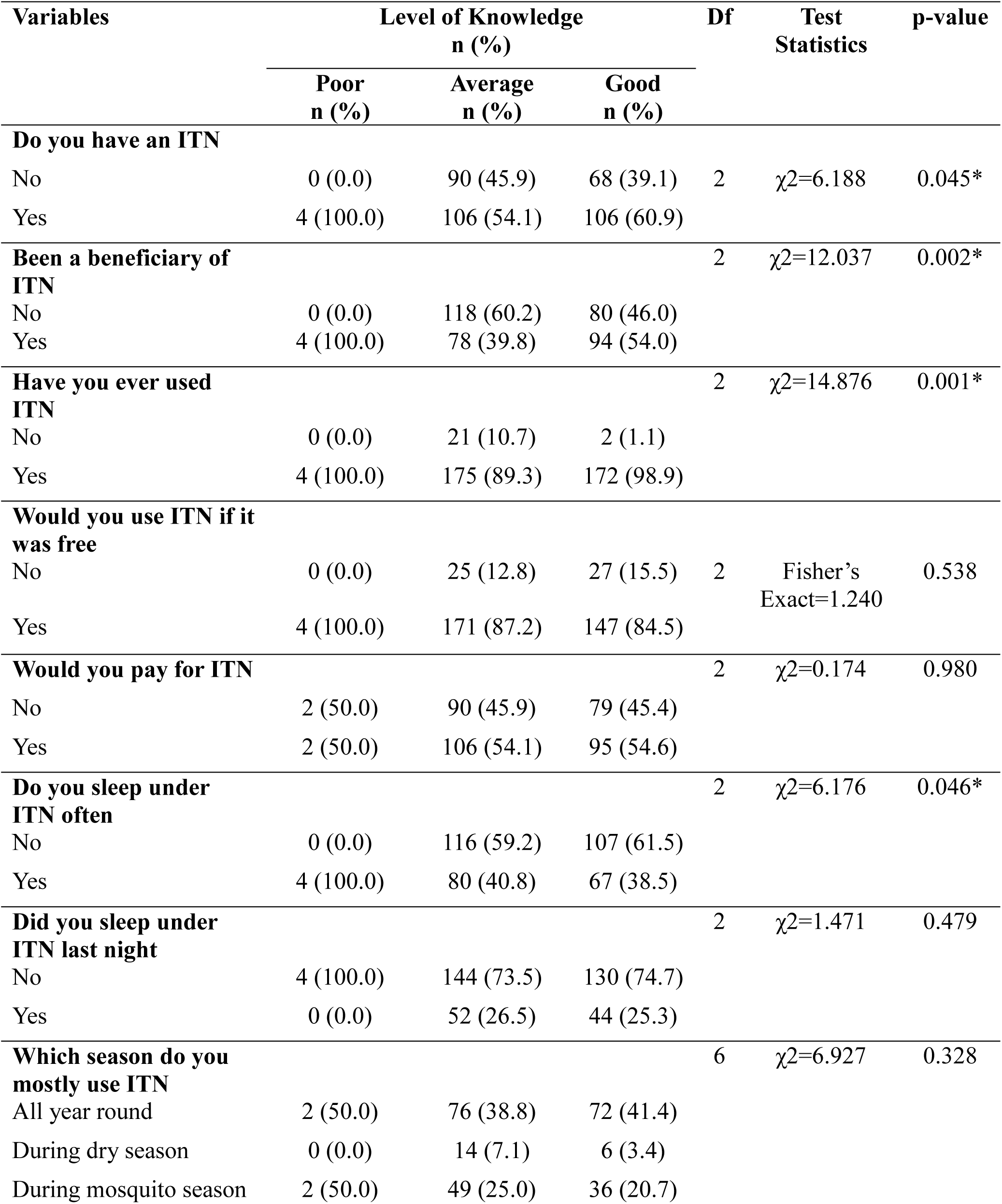
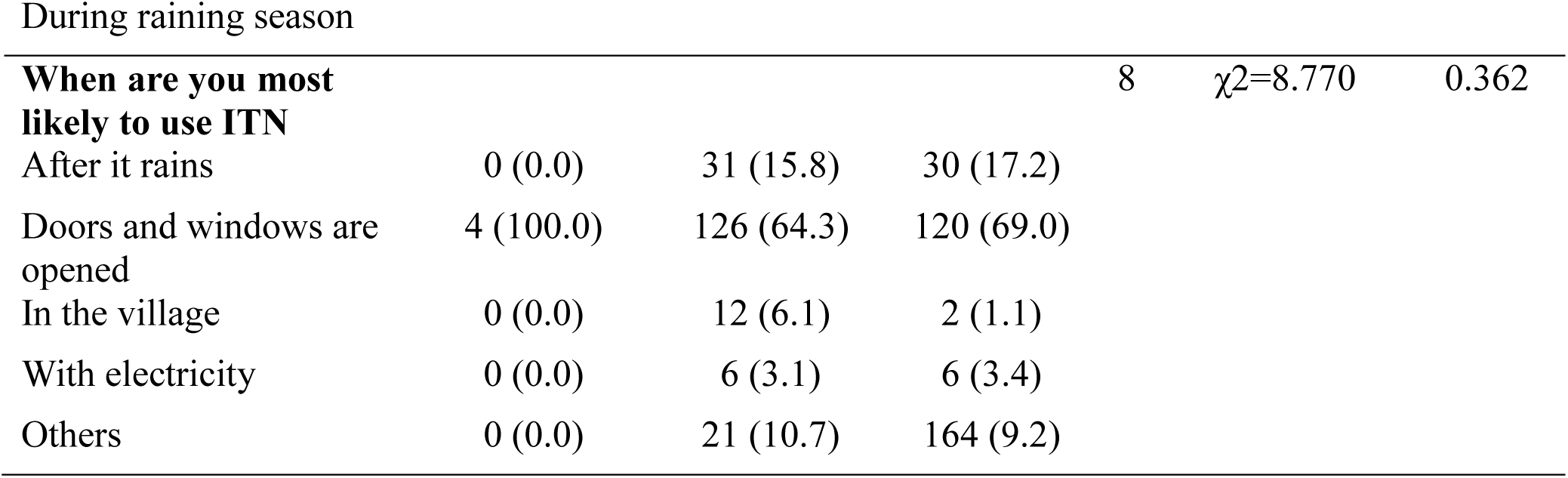
RELATIONSHIP BETWEEN LEVEL OF KNOWLEDGE AND ATTITUDE TOWARDS USE OF ITN.

| Variables | Level of Knowledge<br>n (%) |  |  | Df | Test<br>Statistics | p-value |
| --- | --- | --- | --- | --- | --- | --- |
|  | Poor<br>n (%) | Average<br>n (%) | Good<br>n (%) |  |  |  |
| **Do you have an ITN** |  |  |  |  |  |  |
| No | 0 (0.0) | 90 (45.9) | 68 (39.1) | 2 | $\chi^2=6.188$ | 0.045* |
| Yes | 4 (100.0) | 106 (54.1) | 106 (60.9) |  |  |  |
| **Been a beneficiary of ITN** | | | | 2 | $\chi^2=12.037$ | 0.002* |
| No | 0 (0.0) | 118 (60.2) | 80 (46.0) |  |  |  |
| Yes | 4 (100.0) | 78 (39.8) | 94 (54.0) |  |  |  |
| **Have you ever used ITN** | | | | 2 | $\chi^2=14.876$ | 0.001* |
| No | 0 (0.0) | 21 (10.7) | 2 (1.1) |  |  |  |
| Yes | 4 (100.0) | 175 (89.3) | 172 (98.9) |  |  |  |
| **Would you use ITN if it was free** |  |  |  |  |  |  |
| No | 0 (0.0) | 25 (12.8) | 27 (15.5) | 2 | Fisher's<br>Exact=1.240 | 0.538 |
| Yes | 4 (100.0) | 171 (87.2) | 147 (84.5) |  |  |  |
| **Would you pay for ITN** | | | | 2 | $\chi^2=0.174$ | 0.980 |
| No | 2 (50.0) | 90 (45.9) | 79 (45.4) |  |  |  |
| Yes | 2 (50.0) | 106 (54.1) | 95 (54.6) |  |  |  |
| **Do you sleep under ITN often** | | | | 2 | $\chi^2=6.176$ | 0.046* |
| No | 0 (0.0) | 116 (59.2) | 107 (61.5) |  |  |  |
| Yes | 4 (100.0) | 80 (40.8) | 67 (38.5) |  |  |  |
| **Did you sleep under ITN last night** | | | | 2 | $\chi^2=1.471$ | 0.479 |
| No | 4 (100.0) | 144 (73.5) | 130 (74.7) |  |  |  |
| Yes | 0 (0.0) | 52 (26.5) | 44 (25.3) |  |  |  |
| **Which season do you mostly use ITN** | | | | 6 | $\chi^2=6.927$ | 0.328 |
| All year round | 2 (50.0) | 76 (38.8) | 72 (41.4) |  |  |  |
| During dry season | 0 (0.0) | 14 (7.1) | 6 (3.4) |  |  |  |
| During mosquito season | 2 (50.0) | 49 (25.0) | 36 (20.7) |  |  |  |
| During raining season |  |  |  |  |  |  |
| **When are you most likely to use ITN** | | | | 8 | $\chi^2=8.770$ | 0.362 |
| After it rains | 0 (0.0) | 31 (15.8) | 30 (17.2) |  |  |  |
| Doors and windows are opened | 4 (100.0) | 126 (64.3) | 120 (69.0) |  |  |  |
| In the village | 0 (0.0) | 12 (6.1) | 2 (1.1) |  |  |  |
| With electricity | 0 (0.0) | 6 (3.1) | 6 (3.4) |  |  |  |
| Others | 0 (0.0) | 21 (10.7) | 164 (9.2) |  |  |  |

**Fig 2.**
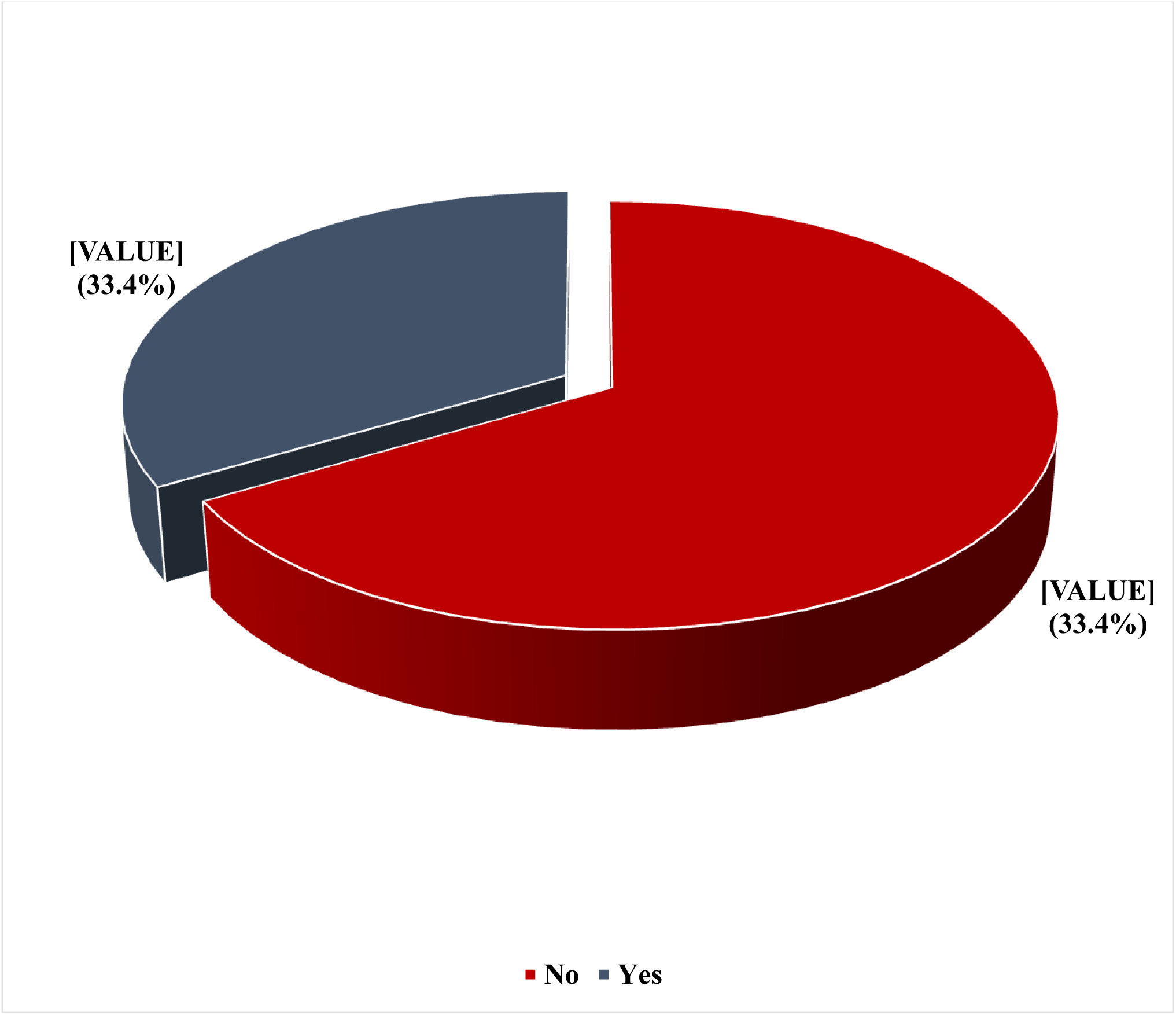
Prevalence of use of insecticide treated nets among participants.

**Table S8.**
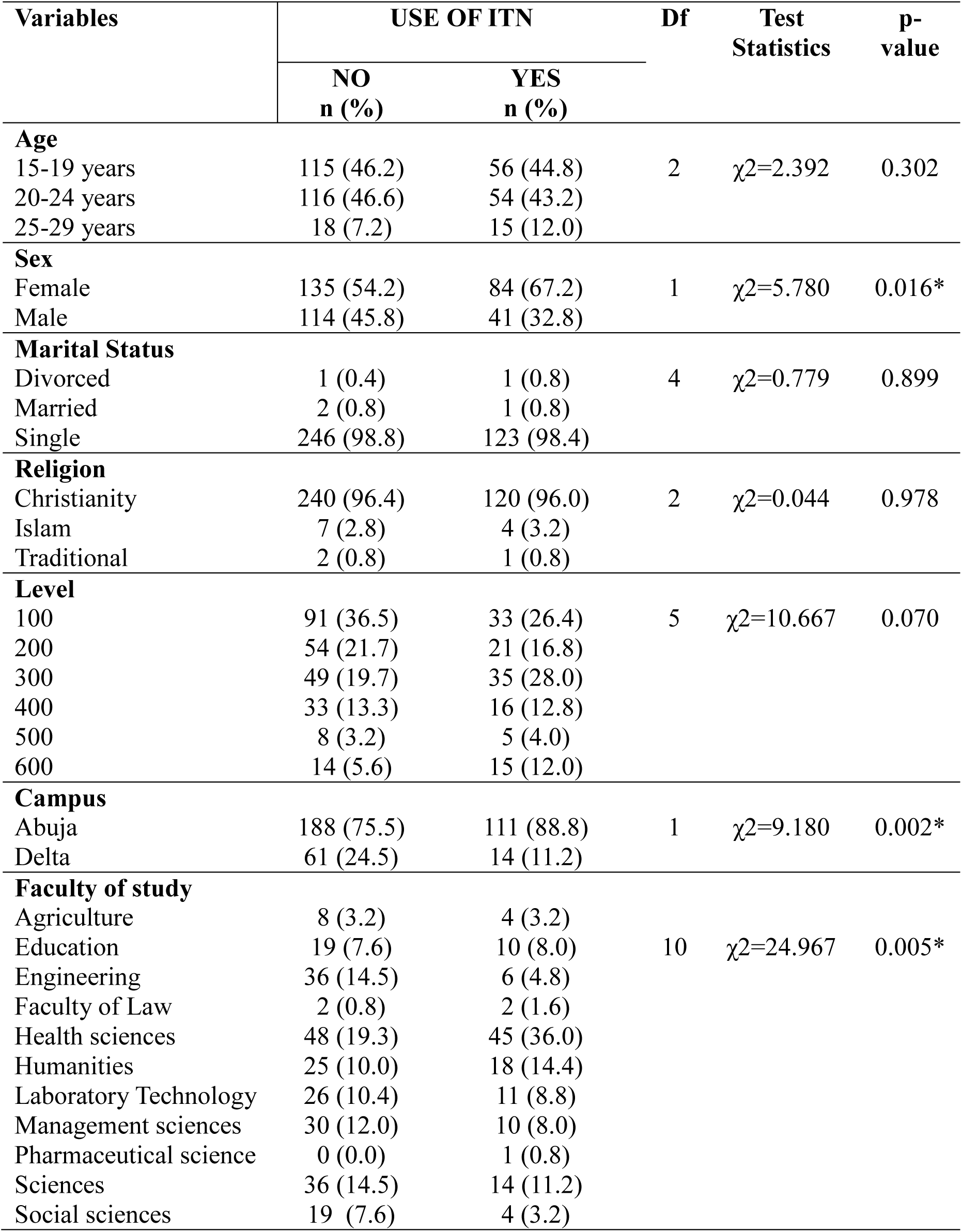
SOCIODEMOGRPAHIC FACTORS ASSOCIATED WITH THE USE OF ITN AMONG HOSTEL RESIDENTS.

